# Multimorbidity in type 1 diabetes is common and associated with increased mortality

**DOI:** 10.1101/2025.09.07.25334819

**Authors:** Anni Ylinen, Stefan Mutter, Stefanie Hägg-Holmberg, Susanna Satuli-Autere, Valma Harjutsalo, Per-Henrik Groop, Lena M Thorn, the FinnDiane Study Group

**Author notes:** Corresponding author: Lena Thorn MD, DMSc, Associate Professor, Department of General Practice and Primary Health Care, University of Helsinki and Helsinki University Hospital, Biomedicum Helsinki, FIN-00014 University of Helsinki, Finland.

## Abstract

**Background:** Multimorbidity in type 1 diabetes has previously not been studied in detail. Therefore, we aimed to assess the prevalence of multimorbidity and its association with mortality in type 1 diabetes.

**Materials and methods:** This observational follow-up study includes 4,069 individuals with type 1 diabetes from the Finnish Diabetic Nephropathy study. The prevalence of multimorbidity (coexistence of two or more chronic conditions) was based on 32 conditions at baseline. Conditions were grouped into three subcategories: vascular comorbidities, autoimmune disorders, and other conditions. Hazard ratios (HR) for all-cause mortality were calculated.

**Results:** The prevalence of multimorbidity was 60.4% and increased with age and especially diabetes duration. Multimorbidity was associated with increased risk of mortality, HR 6.0 (95% CI 4.6–7.8), p<0.001. The HR for mortality increased by each additional condition and was 37.9 (95% CI 25.7–56.0) in those with ≥ eight conditions. Vascular comorbidities and other conditions were associated with increased mortality, HRs 5.9 (4.4–7.9) and 3.8 (2.4–5.9), p<0.001, separately, and in combination, HR 11.2 (8.3–15.2), p<0.001. Autoimmune disorders did not influence mortality.

**Conclusions:** Multimorbidity in type 1 diabetes is common and is associated with increased mortality. Comprehensive evaluation of all additional conditions is needed to tailor treatment individually.

**Key Messages:**

- The prevalence of multimorbidity in our study was 60.4% already at a median age of 38 years and the more chronic conditions an individual had, the higher the risk of mortality.
- A holistic approach is needed in the treatment of individuals with type 1 diabetes, considering also other conditions that negatively impact prognosis.

## Introduction

Lately, the interest in multimorbidity has grown due to its substantial effects on individuals, healthcare, and society (1). Multimorbidity is commonly defined as the coexistence of two or more chronic conditions. It is often confused with comorbidity, which refers to additional disorders in relation to one index disease, whereas multimorbidity addresses all conditions in one individual at a certain point of time, not necessarily with any connection to each other (2). In the general population, the prevalence of multimorbidity increases with age (3), and is associated with increased mortality and decreased quality of life (4).

A rise in multimorbidity has been seen in recent years due to more effective treatments, leading to individuals living longer (5,6). Diabetes is associated with higher probability of other chronic conditions (7). Type 1 diabetes is often followed by other chronic conditions at a relatively young age. It is typically associated with other autoimmune diseases (8,9) as well as micro- and macrovascular comorbidities. Furthermore, individuals with type 1 diabetes may have other conditions, such as cancer or dementia, with even more significant effect on their prognosis (10,11). Additional conditions may also influence the treatment of their diabetes, for example depression and dementia profoundly impact self-care (12).

Despite this, multimorbidity in type 1 diabetes has gained little attention. An American study found that multimorbidity occurs in 84% of hospitalized adults with type 1 diabetes (13). Furthermore, the study showed that the number of chronic conditions relates to a higher probability of depression (13). The most common disorder to coincide with type 1 diabetes has been found to be hypertension (13,14), followed by dyslipidemia, dorsalgia, diabetic neuropathy, and depression (14). In addition, multimorbidity in type 1 diabetes has been associated with severe hypoglycemic episodes (15,16).

No unified measure of multimorbidity is yet available. Different measures have been created (e.g., the Charlson Comorbidity Index (17), the Cumulative Illness Rating Scale (18), the Index of Coexistent Disease (19), and the Elixhauser Index (20)) and simple counts of diseases have also been widely adopted. The choice of measure used to define multimorbidity is affected by the available data and targeted study outcomes. Disease counts have for example been recommended, when multiple outcomes are being considered and in the absence of a validated measure. (21) In 2022, a Delphi consensus study was published with recommendations by a panel of international experts on the definition and measurement of multimorbidity in research (22). The professional and public panel concluded on a list of 24 conditions to always be included when studying multimorbidity, and 37 conditions to usually be included.

Recently, multimorbidity in type 1 diabetes was identified as a knowledge gap (1). Therefore, our aim was to explore the prevalence of multimorbidity and the number of chronic conditions according to sex, age, diabetes duration, and age at onset of type 1 diabetes. Further, we aimed to assess the impact of multimorbidity on mortality. In addition, we aimed to separate the risk related to autoimmune diseases, vascular comorbidities, and other chronic conditions.

## 2. Material and Methods

### 2.1 Study population

Our study is part of the Finnish Diabetic Nephropathy (FinnDiane) Study, which is an ongoing, nationwide multicenter study founded in 1997, including 77 study centers (Supplemental Table S1) (23). The aim of the FinnDiane Study is to identify clinical, environmental, and genetic risk factors for diabetic complications. The final goal is to prevent diabetic complications, to enable personalized treatment, and to improve the quality of life of people with diabetes. Out of the 4,961 individuals with type 1 diabetes, who participated in the FinnDiane Study before the end of 2017, we excluded 892 with incomplete data on the selected conditions. The characteristics of the included vs the excluded participants are provided in Supplemental Table S2, with no observed difference in sex, age, diabetes duration, or age at diabetes onset. The cohort is nationally representative and consists of 4,069 individuals. Sex was defined as the sex assigned at birth. Type 1 diabetes was defined as age at onset below 40 years and insulin therapy initiated within one year from diagnosis. The FinnDiane Study does not register the ethnicity of the participants, but the vast majority are of Caucasian origin.

The study has been approved by the Ethics Committee of the Helsinki and Uusimaa Hospital District. Written informed consent was given by each participating individual, and the study was conducted in accordance with the Declaration of Helsinki.

### 2.2 Baseline study visit

Data at baseline were collected at regular visits to the attending physician, where information on medical history and medications was registered. Height and body weight, and office blood pressure were measured. Blood samples were drawn and analyzed for blood lipids and lipoproteins, creatinine, and HbA_1c_. In addition, 24-hour urine samples were collected to measure the urinary albumin excretion. The participants completed questionnaires regarding their medical history and smoking habits.

### 2.3 Definition of multimorbidity

Multimorbidity was defined based on the individuals having diabetes and at least one more chronic condition. The burden of multimorbidity was defined according to the number of conditions. The accumulation of diseases was defined based on the number of chronic conditions at baseline from a list consisting of 32 conditions (Supplemental Table S3). This list was gathered after reviewing the literature; the conditions were chosen based on a previous large study and the Charlson Index (3,17), and we included all chronic conditions with available data. The included conditions show a good overlap with the Delphi consensus study, that was published after our data collection (22).

Data were collected from clinical records, questionnaires filled out both by the individual and the attending physician, the Finnish Care Register for Health Care, and Statistics Finland. Autoimmune diseases were also identified from the Finnish National Drug Reimbursement Register, the Drug Prescription Register, and the National Health Insurance Dietary Grant Register (8). Data from all different sources were combined. Information on definition of conditions is provided in Supplemental Table S3. The conditions were further grouped into three subcategories: vascular comorbidities, autoimmune diseases, and other disorders (Table 1). For sub-analysis, the cohort was also grouped by age at onset below (n=2,240) or above 15 years (n=1,829).

**Table 1.** Clinical characteristics of the cohort.

| **Variable** | **All data (n=4,069)** |
| --- | --- |
| Women, % (n) | 48.6 (1,997) |
| Age, years | 37.7 (28.9–46.9) |
| Diabetes duration, years | 21.8 (12.2–31.4) |
| Age at diabetes onset, years | 13.9 (9.1–22.1) |
| BMI, kg/m<sup>2</sup> | 25.0 ± 3.6 |
| Systolic blood pressure, mmHg | 132 (121–145) |
| Diastolic blood pressure, mmHg | 80 (72–86) |
| Total cholesterol, mmol/l | 4.92 ± 0.98 |
| LDL cholesterol, mmol/l | 3.04 ± 0.89 |
| HDL cholesterol, mmol/l | 1.34 ± 0.39 |
| Triglycerides, mmol/l | 1.03 (0.77–1.48) |
| HbA<sub>1c</sub>, % (mmol/mol) | 8.4 ± 1.5 (68 ± 16) |
| History of smoking, % (n) | 46.1 (1,876) |

Data are presented as mean ± SD, median (IQR), or % (n)

### 2.4 Follow-up data on mortality

Data on mortality were retrieved from the Finnish Cause of Death Register, Statistics Finland, until the end of 2017. Follow-up data were available for all participants. Follow-up started at the FinnDiane baseline visit and continued until death or the end of 2017, with median follow-up of 16.7 (13.6–18.8) years.

### 2.5 Statistical analyses

Continuous variables were tested for normality. Parametric continuous variables were tested with the unpaired t test to compare means and non-parametric ones were tested with Mann-Whitney U-tests. The results are presented as means with standard deviation or medians with interquartile ranges. The χ^2^-test was used to test differences in categorical variables between groups and the Fisher’s exact test was used if observations in groups were ≤ 5.

Hazard ratios (HRs) for all-cause mortality were calculated in sex- and age-adjusted Cox regression models, according to the number of diseases, the three subcategories, and the different conditions separately. The results are presented as HR with 95% confidence interval (CI).

A p-value <0.05 was considered statistically significant. Statistical analyses were performed with SPSS Statistics 29.0 software (IBM Corporation, Armonk, NY, USA).

## 3. Results

### 3.1 Baseline characteristics of the cohort

The cohort consists of 4,069 individuals with type 1 diabetes (48.6% women). Median age at baseline was 37.7 (28.9–46.9) years, median duration of diabetes was 21.8 (12.2–31.4) years, and median age at onset was 13.9 (9.1–22.1) years. Clinical characteristics of the cohort are presented in Table 1.

### 3.2 Prevalence of multimorbidity and chronic conditions

The prevalence of multimorbidity was 60.4 % (n=2,458). The prevalence of the different conditions is presented in Supplemental Table S4. Hypertension was the most common chronic condition (40.6%, n=1,653), followed by severe retinopathy (35.1%, n=1,427), diabetic kidney disease (24.6%, n=1,002), thyroid disease (8.8%, n=358), and coronary heart disease (5.9%, n=242).

The most common subcategory was vascular comorbidities, present in 49.2%, (n=2,000), autoimmune disorders were present in 12.7% (n=515), and other conditions in 19.8% (n=807). The prevalence of number of conditions in the participants is presented in Table 2.

**Table 2.** Prevalence and risk of mortality according to number of diseases.

| **Number of diseases** | **Prevalence % (n)** | **Hazard ratio (HR), age and sex adjusted** |
| --- | --- | --- |
| 1 (only diabetes) | 39.6 (1,611) | Reference |
| 2 | 20.1 (818) | 1.7 (1.2–2.4)* |
| 3 | 12.7 (518) | 4.2 (3.0–5.8)* |
| 4 | 12.7 (518) | 8.4 (6.2–11.3)* |
| 5 | 6.8 (276) | 13.0 (9.5–17.8)* |
| 6 | 3.6 (147) | 18.4 (13.0–26.1)* |
| 7 | 2.5 (103) | 29.0 (20.3–41.6)* |
| 8 or more | 1.9 (78) | 37.9 (25.7–56.0)* |

Data are hazard ratios with 95% confidence intervals. \*p≤0.005

### 3.3 Multimorbidity by sex

There was no difference in the prevalence of multimorbidity between men and women (60.8% [n=1,271] *vs* 60.0% [n=1,187] p=0.641). Of the subcategories, vascular comorbidities were more common in men (53.1% [n=1,110] *vs* 45.0% [n=890], p<0.001), whereas autoimmune disorders were more frequent in women (18.2% [n=360] *vs* 7.4% [n=155], p<0.001). There was no difference between men and women regarding the prevalence of the other conditions (18.9% [n=396] *vs* 20.8% [n=411], p=0.137).

### 3.4 Multimorbidity by age

The prevalence of multimorbidity clearly increased by age (Figure 1A): the prevalence in individuals <25 years was 25.7% (n=170), 25–30 years 38.6% (n=181), 30–35 years 54.7% (n=327), 35–40 years 65.2% (n=368), 40–45 years 72.2% (n=416), 45–50 years 78.0% (n=369), 50–55 years 84.0% (n=336), 55–60 years 85.0% (n=153), and >60 years 93.2% (n=138), p<0.001. The figure also displays the number of conditions in each age group.

**Figure 1.**
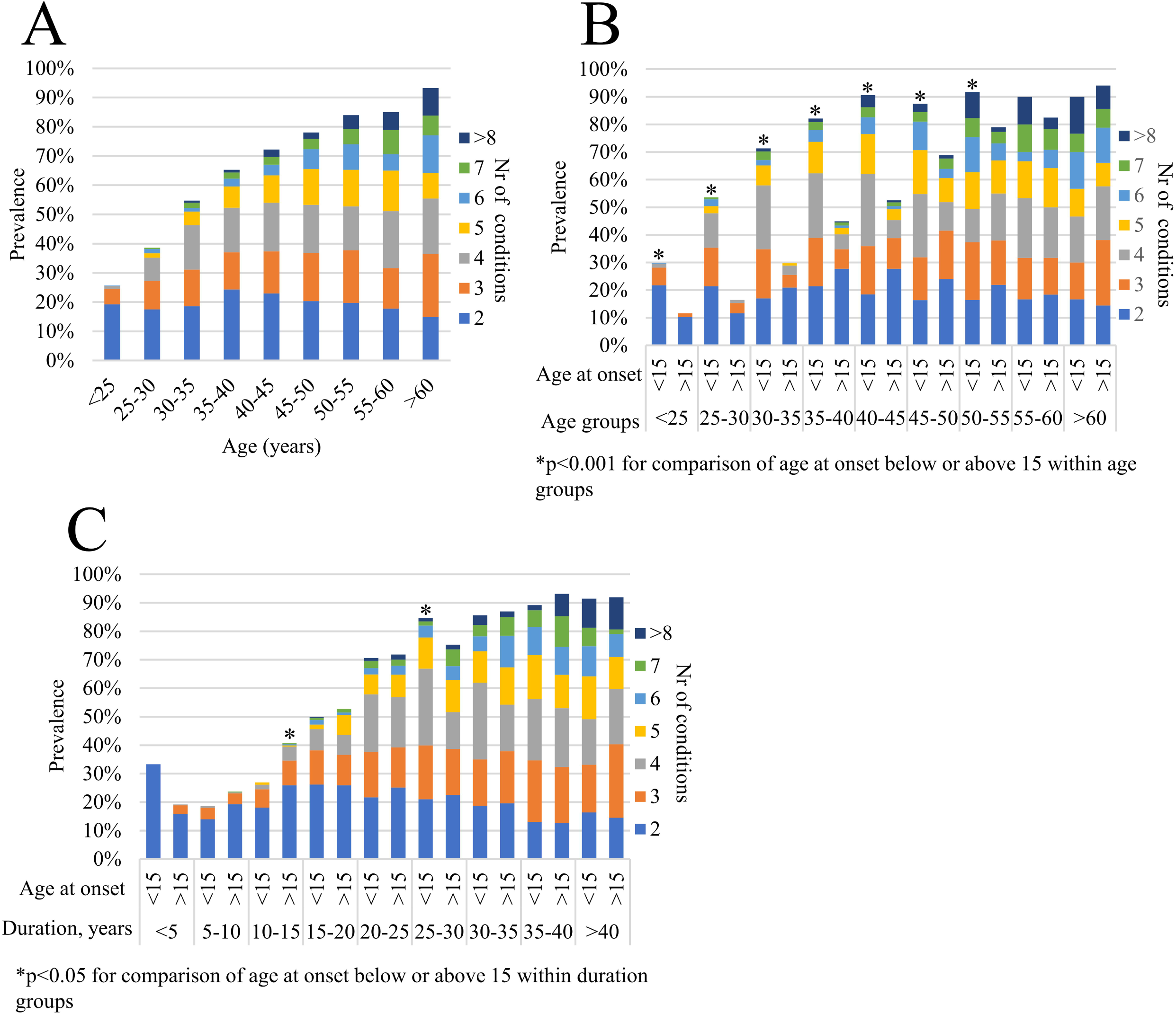
Prevalence of multimorbidity and the number of conditions. Panel A: According to age groups, Panel B: According to age at onset and age groups, and Panel C: According to age at onset and diabetes duration.

### 3.5 Multimorbidity by age at onset of diabetes

All participants were divided into two groups based on their age at onset of diabetes, below or above 15 years. Individuals with age at onset of diabetes below 15 years were younger (34.4 [25.5–43.6] *vs* 41.6 [32.8–50.6] years, p<0.001), had a longer duration of diabetes (26.0 [17.2–34.4] *vs* 16.4 [7.4–26.2] years, p<0.001), and by default a lower age at onset (9.6 [6.1– 12.3] *vs* 23.3 [18.6–29.0] years, p<0.001). The prevalence of multimorbidity was significantly higher in individuals with age at onset of diabetes below 15 years (67.5% [n=1,511] *vs* 51.8% [n=947], p<0.001), despite the younger age. The prevalence of multimorbidity, as well as the number of conditions according to age at onset below or above 15 years, in the different age groups is presented in Figure 1B. The prevalence of multimorbidity was clearly higher in the onset <15 years group until the age of 55, after which there was no significant difference between the two groups. Further, the prevalence of multimorbidity in groups by duration of diabetes, according to age at onset below or above 15 years, is presented in Figure 1C. This figure shows no clear trend in the differences in the prevalence of multimorbidity between the age at onset groups. In all except for two duration-categories, there was no significant difference in the prevalence of multimorbidity between individuals with diabetes onset below or above 15 years.

### 3.6 Multimorbidity and mortality

During a median follow-up time of 16.7 (13.6–18.8) years, 784 (19.3%) of the participants died. In age- and sex-adjusted Cox-regression analysis, multimorbidity was associated with increased risk of mortality, HR 6.01 (4.55–7.94, p<0.001). Sex- and age-adjusted Cox-regression analysis was performed according to the number of diseases and each higher number of diseases was associated with significantly higher risk of mortality (Table 2). The HRs for mortality for the separate conditions are presented in Supplemental Table S4.

Figure 2 displays the HRs for mortality from age- and sex-adjusted Cox-regression analyses in the different subcategories. Vascular comorbidities (p<0.001) and other conditions (p<0.001) alone were associated with increased risk of mortality, while autoimmune disorders alone were not associated with any increased risk of mortality. Furthermore, in individuals with both vascular comorbidities and other conditions, the risk of mortality was markedly increased. Autoimmune disorders did not further increase the risk of mortality in combination with vascular comorbidities or other conditions, or when all three subcategories were combined.

**Figure 2.**
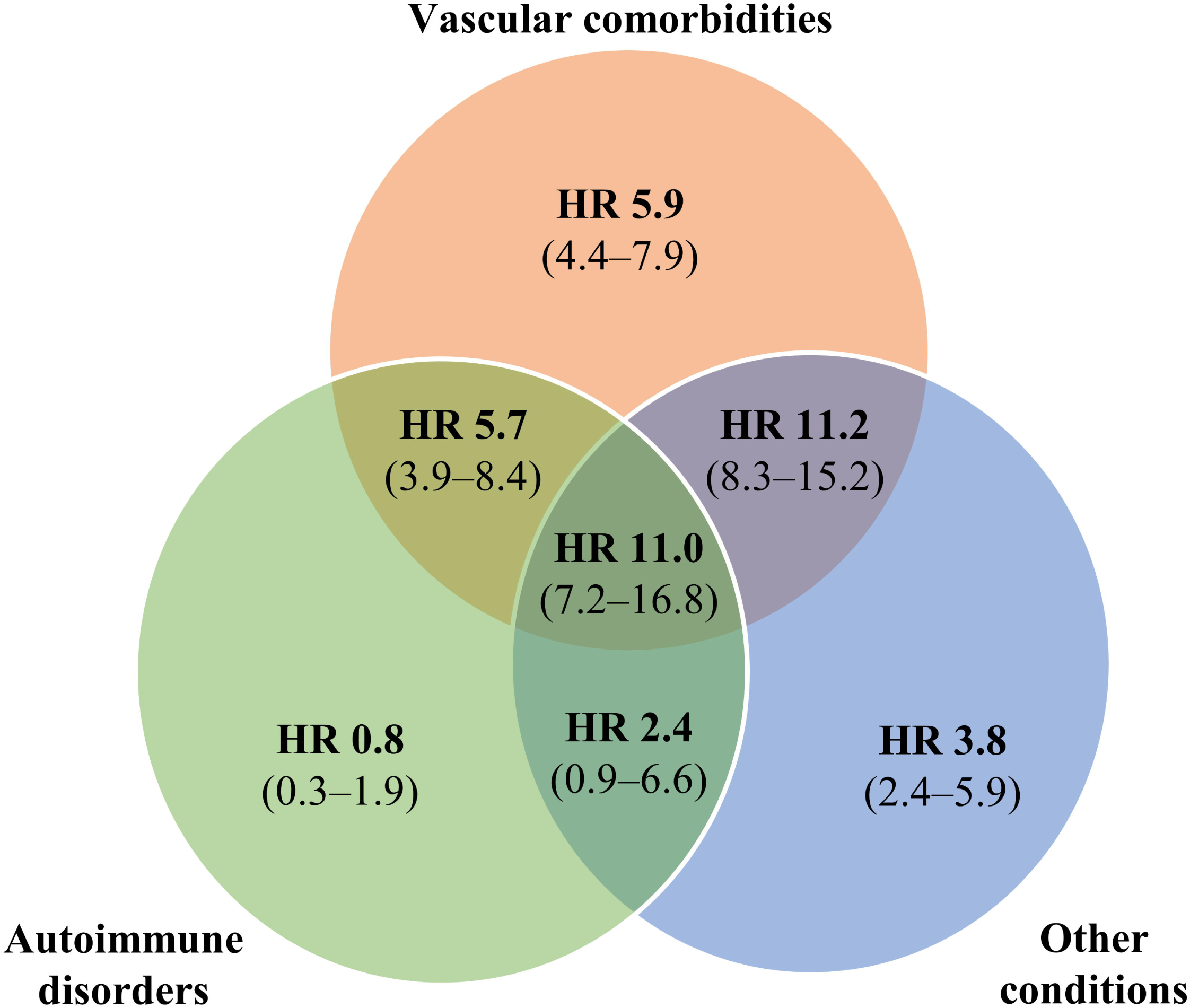
Risk of mortality according to subcategories of the different conditions.

## 4. Discussion

We found that the prevalence of multimorbidity in type 1 diabetes is high and increases by age and especially by duration of type 1 diabetes. Multimorbidity is already present at young age, most clearly in individuals with diabetes onset before the age of 15 years. Multimorbidity in type 1 diabetes is associated with increased mortality: the higher the number of conditions, the higher the risk of mortality, with a striking 38-fold increased risk of mortality in those with eight chronic conditions or more. The increased mortality related to multimorbidity is largely driven by vascular comorbidities, and this risk is further increased by other chronic conditions, while autoimmune disorders do not influence the risk. These findings help us understand the previously declared research gap in multimorbidity in type 1 diabetes (1).

In our study, the prevalence of multimorbidity in type 1 diabetes was high, 60.4%, as we expected. Previous studies have found the prevalence to be even higher, 82–84% (13,14). The individuals in these studies were, however, older and one of the studies (13) was conducted on hospitalized patients, which likely included sicker individuals, and this could have impacted the higher prevalence of multimorbidity. The number and type of studied conditions can also impact the reported prevalence. We chose to focus on more severe chronic conditions than one of the previous studies (14), which also included milder diseases. Most striking in our study is that our participants are relatively young, and in relation to this, the prevalence of multimorbidity is significant. In line with previous studies, we found that hypertension was the most common additional condition in type 1 diabetes (13,14).

While multimorbidity in type 1 diabetes has been understudied previously, in type 2 diabetes, the health burden has been shown to be markedly greater than in individuals without diabetes (24). The prevalence of multimorbidity in type 2 diabetes has generally been studied on older individuals than in our study, and the prevalence has been reported to be 70–93% in individuals with mean age of 60–71 years (25–28). Individuals with youth-onset type 2 diabetes have been shown to have a high burden of microvascular complications (29).

Our study shows that in type 1 diabetes, the high prevalence of multimorbidity already at young age is mainly present in individuals with young age at onset, below 15 years. Interestingly, when comparing these groups (age at onset of diabetes below or above 15 years) at the same duration of diabetes, the prevalence of multimorbidity was similar between the groups, indicating that the duration of diabetes is the factor driving multimorbidity, rather than the young age at onset *per se*.

We showed that multimorbidity in type 1 diabetes is associated with increased risk of mortality and that the risk is notably greater with increasing multimorbidity. This has not been studied in type 1 diabetes previously, but multimorbidity has been associated with increased risk of mortality in type 2 diabetes (25). We also showed that the increased mortality is largely driven by vascular complications, with a six-fold increase in risk of mortality. This finding was expected based on previous research (30–32). Interestingly, the other conditions also had a substantial impact on mortality with an almost four-fold increase and was especially seen in combination with vascular comorbidities, leading to an 11-fold increase in mortality. Furthermore, autoimmune diseases were not associated with increased mortality alone, nor did they affect the risk related to vascular complications or other conditions. A recent Swedish study found similar results, with no link between autoimmune diseases and mortality risk (33).

Studying multimorbidity is complicated, partly because widely various measures have been used (34). Recently researchers have published a Delphi Consensus Study for guidance to promote consistency in studying multimorbidity and for future studies to be more comparable (22). However, during the process of deciding which conditions to include in studies, it is impossible to include all the conditions in any individual. Many conditions remain undiagnosed, and some are difficult to include in studies on multimorbidity due to how they are reported. Consequently, our studied prevalence of multimorbidity is more likely an underestimation of the actual prevalence of multimorbidity in type 1 diabetes.

This study represents a well-characterized cohort of individuals with type 1 diabetes, with a long follow-up time, which is a major strength of the study. However, we have likely not been able to detect all chronic conditions. Another limitation of this study is that the definition of some conditions may be imprecise, for example the conditions defined based on the use of medications. Often medications have more than one indication, thus, they can be used for more than one disease. Despite the medications not being specific, the use of a medication reveals something about the health of an individual and, in addition, describes the health burden, which is what we aimed to capture in this study. It is important to bear in mind that our study is a good representation of individuals of Caucasian origin, but our results may not be generalizable to other populations.

In conclusion, multimorbidity in type 1 diabetes is very common at young age and the prevalence increases by age, and especially by longer diabetes duration. Our findings highlight the importance of disease prevention in type 1 diabetes, since multimorbidity is associated with increased risk of mortality. In a clinical setting, our study is a reminder that people with type 1 diabetes already at young age present with many other conditions that impact their prognosis, and it is, thus, important to consider the whole disease burden of an individual, in addition to the separate conditions. The impact of multimorbidity on individuals with type 1 diabetes requires further research to understand the interplay between different conditions and their significance for the management of diabetes.

## Supporting information

Online-Only Supplemental Material

## Data Availability

The datasets generated and/or analyzed during the current study are not publicly available due to the local legislation and the written consents of the FinnDiane study participants, which do not allow sharing individual-level phenotype data. The data that support the findings are available from the corresponding author upon reasonable request.

## Abbreviations

FinnDiane: Finnish Diabetic Nephropathy Study

## Acknowledgements

Personal thanks: We acknowledge all the participants and the physicians and nurses at each study center participating in the collection of the study population (Supplemental Table S1). We are indebted to the late Carol Forsblom, the international coordinator of the FinnDiane Study Group, for his contribution to this work.

## Author Contributions and Guarantor Statement

A.Y. had the main responsibility for analyzing the data and writing the first draft of the paper and contributed to the acquisition and processing of data. As a supervisor, L.M.T. greatly contributed to study design, acquisition and processing of data, data analysis, and critical revision of the paper. S.M. contributed to study design, data analysis, and critical revision of the paper. S.H.-H., S.S.-A., V.H., and P.-H.G contributed to study design, acquisition of data, and critical revision of the paper. All authors reviewed and approved the final version of the manuscript. L.M.T. is the guarantor of this work and, as such, had full access to all the data and takes responsibility for the integrity of the data and the accuracy of the data analysis.

