## Supplementary material for "Multimorbidity in type 1 diabetes is common and associated with increased mortality": Online-Only Supplemental Material

Supplemental Table S1. The Finnish Diabetic Nephropathy Study Centers

| FinnDiane Study Centers | Physicians and nurses |
| --- | --- |
| Anjalankoski Health Center | S.Koivula, T.Uggeldahl |
| Central Finland Central Hospital, Jyväskylä | T.Forslund, A.Halonen, A.Koistinen, P.Koskiaho, M.Laukkanen, J.Saltevo, M.Tiihonen |
| Central Hospital of Åland Islands, Mariehamn | M.Forsen, H.Granlund, A.-C.Jonsson, B.Nyroos |
| Central Hospital of Kanta-Häme, Hämeenlinna | P.Kinnunen, A.Orvola, T.Salonen, A.Vähänen |
| Central Hospital of Kymenlaakso, Kotka | R.Paldanius, M.Riihelä, L.Ryysy |
| Central Hospital of Länsi-Pohja, Kemi | H.Laukkanen, P.Nyländen, A.Sademies |
| Central Ostrobothnian Hospital District, Kokkola | S.Anderson, B.Asplund, U.Byskata, P.Liedes, M.Kuusela, T.Virkkala |
| City of Espoo Health Center: |  |
| Espoonlahti | A.Nikkola, E.Ritola |
| Tapiola | M.Niska, H.Saarinen |
| Samaria | E.Oukko-Ruponen, T.Virtanen |
| Viherlaakso | A.Lyytinen |
| City of Helsinki Health Center: |  |
| Puistola | H.Kari, T.Simonen |
| Suutarila | A.Kaprio, J.Kärkkäinen, B.Rantaeskola |
| Töölö | P.Kääriäinen, J.Haaga, A-L.Pietiläinen |
| City of Hyvinkää Health Center | S.Klemetti, T.Nyandoto, E.Rontu, S.Satuli-Autere |
| City of Vantaa Health Center: |  |
| Korso | R.Toivonen, H.Virtanen |
| Länsimäki | R.Ahonen, M.Ivaska-Suomela, A.Jauhiainen |
| Martinlaakso | M.Laine, T.Pellonpää, R.Puranen |
| Myyrmäki | A.Airas, J.Laakso, K.Rautavaara |
| Rekola | M.Erola, E.Jatkola |
| Tikkurila | R.Lönnblad, A.Malm, J.Mäkelä, E.Rautamo |
| Heinola Health Center | P.Hentunen, J.Lagerstam |
| Helsinki University Hospital, Department of Medicine, Division of Nephrology | T.Claesson, A.Dufva, N.Elonen, M.Eriksson, J.Fagerudd, M.Feodoroff, D.Gordin, P.-H.Groop, O.Heikkilä, K.Hietala, S.Hägg-Holmberg, F.Jansson Sigfrids, M.Korolainen, J.Kytö, S.Lindh, H.Paajanen, K.Pettersson-Fernholm, K.Rimpeläinen, M.Rosengård-Bärlund, M.Rönnback, L.Salovaara, A.Sandelin, M.Saraheimo, S.Satuli-Autere, R.Simonsen, P.Smidtslund, L.Thorn, H.Tikkanen, J.Tuomikangas, A.Tynjälä, K.Uljala, T.Vesisenaho, J.Wadén, A.Ylinen |
| Herttoniemi Hospital, Helsinki | V.Sipilä |
| Hospital of Lounais-Häme, Forssa | T.Kalliomäki, J.Koskelainen, R.Nikkanen, N.Savolainen, H.Sulonen, E.Valtonen |
| Hyvinkää Hospital | L. Norvio, A.Hämäläinen |
| Iisalmi Hospital | E.Toivanen |
| Jokilaakso Hospital, Jämsä | A.Parta, I.Pirttiniemi |
| Jorvi Hospital, Helsinki University Central Hospital | S.Aranko, S.Ervasti, R.Kauppinen-Mäkelin, A.Kuusisto, T.Leppälä, K.Nikkilä, L.Pekkonen |
| Jyväskylä Health Center, Kyllö | K.Nuorva, M.Tiihonen |
| Kainuu Central Hospital, Kajaani | S.Jokelainen, K.Kananen, M.Karjalainen, P.Kemppainen, A-M.Mankinen, A.Reponen, M.Sankari |
| Kerava Health Center | H.Stuckey, P.Suominen |
| Kirkkonummi Health Center | A.Lappalainen, M.Liimatainen, J.Santaholma |
| Kivelä Hospital, Helsinki | A.Aimolahti, E.Huovinen |
| Koskela Hospital, Helsinki | V.Ilkka, M.Lehtimäki |
| Kotka Health Center | E.Pälikkö-Kontinen, A.Vanhanen |
| Kouvola Health Center | E.Koskinen, T.Siitonen |
| Kuopio University Hospital | E.Huttunen, R.Ikäheimo, P.Karhapää, P.Kekäläinen, M.Laakso, T.Lakka, E.Lampainen, L.Moilanen, S. Tanskanen, L.Niskanen, U.Tuovinen, I.Vauhkonen, E.Voutilainen |
| Kuusamo Health Center | T.Kääriäinen, E.Isopoussu |
| Kuusankoski Hospital | E.Kilkki, I.Koskinen, L.Riihelä |
| Laakso Hospital, Helsinki | T.Meriläinen, P.Poukka, R.Savolainen, N.Uhlenius |
| Lahti City Hospital | A.Mäkelä, M.Tanner |
| Lapland Central Hospital, Rovaniemi | L.Hyvärinen, K.Lampela, S.Pöykkö, T.Rompasaari, S.Severinkangas, T.Tulokas |
| Lappeenranta Health Center | P. Erola, L.Härkönen, P.Linkola, T.Pekkanen, I.Pulli, E.Repo |
| Lohja Hospital | T.Granlund, K.Hietanen, M.Porrassalmi, M.Saari, T.Salonen, M.Tiikkainen, |
| Länsi-Uusimaa Hospital, Tammisaari | I.-M.Jousmaa, J.Rinne |
| Loimaa Health Center | A.Mäkelä, P.Eloranta |
| Malmi Hospital, Helsinki | H.Lanki, S.Moilanen, M.Tilly-Kiesi |
| Mikkeli Central Hospital | A.Gynther, R.Manninen, P.Nironen, M.Salminen, T.Vänttinen |
| Mänttä Regional Hospital | I.Pirttiniemi, A-M.Hänninen |
| North Karelian Hospital, Joensuu | U-M.Henttula, P.Kekäläinen, M.Pietarinen, A.Rissanen, M.Voutilainen |
| Nurmijärvi Health Center | A.Burgos, K.Urtamo |
| Oulaskangas Hospital, Oulainen | E.Jokelainen, P-L.Jylkkä, E.Kaarlela, J.Vuolaspuro |
| Oulu Health Center | L.Hiltunen, R.Häkkinen, S.Keinänen-Kiukaanniemi |
| Oulu University Hospital | R.Ikäheimo |
| Päijät-Häme Central Hospital | H.Haapamäki, A.Helanterä, S.Hämäläinen, V.Ilvesmäki, H.Miettinen |
| Palokka Health Center | P.Sopanen, L.Welling |
| Pieksämäki Hospital | V.Sevtsenko, M.Tamminen |
| Pietarsaari Hospital | M-L.Holmbäck, B.Isomaa, L.Sarelin |
| Pori City Hospital | P.Ahonen, P.Merisalo, E.Muurinen, K.Sävelä |
| Porvoo Hospital | M.Kallio, B.Rask, S.Rämö |
| Raahe Hospital | A.Holma, M.Honkala, A.Tuomivaara, R.Vainionpää |
| Rauma Hospital | K.Laine, K.Saarinen, T.Salminen |
| Riihimäki Hospital | P.Aalto, E.Immonen, L.Juurinen |
| Salo Hospital | A.Alanko, J.Lapinleimu, P.Rautio, M.Virtanen |
| Satakunta Central Hospital, Pori | M.Asola, M.Juhola, P.Kunelius, M.-L.Lahdenmäki, P.Pääkkönen, M.Rautavirta |
| Savonlinna Central Hospital | T.Pulli, P.Sallinen, M.Taskinen, E.Tolvanen, T.Tuominen, H.Valtonen, A.Vartia, S-L.Viitanen |
| Seinäjoki Central Hospital | O.Antila, E.Korpi-Hyövälti, T.Latvala, E.Leijala, T.Leikkari, M.Punkari N.Rantamäki, H.Vähävuori |
| South Karelia Central Hospital, Lappeenranta | T.Ensala, E.Hussi, R.Härkönen, U.Nyholm, J.Toivanen |
| Tampere Health Center | A.Vaden, P.Alarotu, E.Kujansuu, H.Kirkkopelto-Jokinen, M.Helin, S.Gummerus, L.Calonius, T.Niskanen, T.Kaitala, T.Vatanen |
| Tampere University Hospital | P. Hannula, I.Ala-Houhala, R.Kannisto, T.Kuningas, P.Lampinen, M.Määttä,H.Oksala, T.Oksanen, A.Putila, H.Saha, K.Salonen, H.Tauriainen, S.Tulokas |
| Tiirismaa Health Center, Hollola | T.Kivelä, L.Petlin, L.Savolainen |
| Turku Health Center | A.Artukka, I.Hämäläinen, L.Lehtinen, E.Pyysalo, H.Virtamo, M.Viinikkala, M.Vähätalo |
| Turku University Central Hospital | K.Breitholz, R.Eskola, K.Metsärinne, U.Pietilä, P.Saarinen, R.Tuominen, S.Äyräpää |
| Vaajakoski Health Center | K.Mäkinen, P.Sopanen |
| Valkeakoski Regional Hospital | S.Ojanen, E.Valtonen, H.Ylönen, M.Rautiainen, T.Immonen |
| Vammala Regional Hospital | I.Isomäki, R.Kroneld, L.Mustaniemi, M.Tapiolinna-Mäkelä |
| Vasa Central Hospital | S.Bergkulla, U.Hautamäki, V-A.Myllyniemi, I.Rusk |

Supplemental Table S2. Clinical characteristics of the cohort compared to the excluded participants without information on all 32 conditions selected for the study.

| **Variable** | **Study cohort (n=4,069)** | **Excluded participants (n=892)** | **p-value** |
| --- | --- | --- | --- |
| Women, % (n) | 48.6 (1,997) | 47.1 (420) | 0.416 |
| Age, years | 37.7 (28.9–46.9) | 37.1 (28.1–47.9) | 0.964 |
| Diabetes duration, years | 21.8 (12.2–31.4) | 22.1 (13.5– 32.6) | 0.083 |
| Age at diabetes onset, years | 13.9 (9.1–22.1) | 13.4 (8.3–20.5) | 0.052 |
| BMI, kg/m^2^ | 24.7 (22.5–27.0) | 24.9 (22.4–27.5) | 0.096 |
| Systolic blood pressure, mmHg | 132 (121–145) | 132 (121–146) | 0.994 |
| Diastolic blood pressure, mmHg | 80 (72–86) | 79 (71–85) | 0.002 |
| Total cholesterol, mmol/l | 4.92 ± 0.98 | 4.77 ± 1.04 | <0.001 |
| LDL cholesterol, mmol/l | 3.04 ± 0.89 | 2.80 ± 0.95 | <0.001 |
| HDL cholesterol, mmol/l | 1.34 ± 0.39 | 1.43 ± 0.43 | <0.001 |
| Triglycerides, mmol/l | 1.03 (0.77–1.48) | 1.10 (0.78–1.51) | 0.158 |
| HbA_1c_, % (mmol/mol) | 8.4 ± 1.5 (68 ± 16) | 8.4 ± 1.5 (68 ± 16) | 0.833 |
| History of smoking, % (n) | 46.7 (1,876) | 42.2 (285) | 0.029 |

Data are presented as mean ± SD, median (IQR) or % (n)

Supplemental Tabel S3. Definition of conditions

| **Condition** | **Definition** |
| --- | --- |
| *Vascular comorbidities* |  |
| Hypertension | Use of antihypertensive medication or physician-reported hypertension |
| Severe retinopathy | History of retinal photocoagulation therapy (physician-reported data) |
| Kidney disease | Laboratory measures (eGFR below 60 ml/min/1.73 m^2^, calculated from serum creatinine based on the Chronic Kidney Disease Epidemiology Collaboration (CKD-EPI) equation), albumin excretion rate ≥200 μg/min or ≥300mg/24h in two out of three urine collections, or kidney replacement therapy |
| Coronary heart disease | Physician-reported history of coronary heart disease, coronary revascularization, or acute myocardial infarction |
| Peripheral vascular disease | Physician-reported history of peripheral revascularization or amputation |
| Myocardial infarction | Physician reported history of myocardial infarction |
| Stroke | Physician reported history of stroke |
| Heart failure | History of hospitalization due to heart failure, based on registry data from the Finnish Care Register for Health Care (ICD-9 code: 428, ICD-10 code: I50) |
| *Autoimmune disorders* |  |
| Thyroid disease | Use of medication for hypo- or hyperthyroidism, self-reported data on hypo- or hyperthyroidism from research questionnaire, and history of hypo- or hyperthyroidism based on registry data from the Finnish Care Register for Health Care (ICD-8: 24400, 24409, 24200, 24209, ICD-9: 2452A, 2458X, 2459X, 2420A and ICD-10: E0380, E0389, E039, E050), Drug Prescription Register (ATC) (H03AA01, H03AA02, H03AA03, H03AA05, H03BB01, H03BA02), and Drug Reimbursement Register (Code 104 and ICD-10 E03, E038, E039) |
| Celiac disease | Self-reported data on celiac disease from research questionnaire, history of celiac disease based on registry data from the Finnish Care Register for Health Care (ICD-9: 5790A, ICD-10: K900) and National Health Insurance Dietary Grant register (reimbursement for gluten-free diet) |
| Rheumatoid arthritis | Self-reported data from research questionnaire |
| Addison’s disease | History of Addison’s disease based on registry data from the Finnish Care Register for Health Care (ICD-8: 25510, ICD-9: 2554A, ICD-10 E271, E272) and Drug Reimbursement Register (Code 105 and ICD E271). |
| Multiple sclerosis | History of multiple sclerosis based on registry data from the Finnish Care Register for Health Care (ICD-8: 340–341, ICD-9: 340–341, ICD-10: G35–37) |
| Atrophic gastritis | History of atrophic gastritis based on registry data from the Finnish Care Register for Health Care (ICD-9: 5351X, ICD-10: K294) |
| Myasthenia gravis | History of myasthenia gravis based on registry data from the Finnish Care Register for Health Care (ICD-8: 73300, ICD-9: 3580, ICD-10: G70) |
| *Other conditions* |  |
| Lung disease | Use of inhaled respiratory medication or self-reported data on asthma from research questionnaire |
| Depression | Use of antidepressive medication or self-reported data on depression from research questionnaire |
| Dyspepsia | Use of medication for dyspepsia |
| Chronic pain | Daily use of any painkillers or irregular use of opioids |
| Alcohol misuse | History of hospitalization due to alcohol-related cause, based on registry data from the Finnish Care Register for Health Care (ICD-8: 291, 303, 57100, 57101, 979–980, E860, E854–856, ICD-9: 291, 303, 3050, 3575, 4255, 5353, 5710-5713, 5770D–5770F, 5771C, 5771D, 5772, 5778, 5779, 7903, 980, E860, ICD-10: E244, F10, G621, G721, I426, K292, K70, K852, K8600–8608, O354, T510, X45, Y90, Y91, Z502, Z714, Z721) |
| Anxiety/insomnia | Use of medication for anxiety or insomnia |
| Epilepsy | Self-reported data on epilepsy from research questionnaire, and history of epilepsy based on registry data from the Finnish Care Register for Health Care (ICD-8: 342, ICD-9: 3320, 3321, 330, ICD-10: G40–41) |
| Cancer | History of cancer based on registry data from the Finnish Care Register for Health Care within five years prior to baseline (ICD-9: 140–239, ICD-10: C00-C99, D00–09, D45, D473) |
| Glaucoma | Use of medication for glaucoma |
| Osteoporosis | Use of medication for osteoporosis |
| Substance misuse | History of hospitalization due to substance-related cause, based on registry data from the Finnish Care Register for Health Care (ICD-8 07003, 304, 979, ICD-9 304, 3052–3059, 965, 967, 969, E9801–9804, ICD-10: F11–16, F18–19, T40, T96, Z715, Z722) |
| Schizophrenia | Use of antipsychotic medication |
| Dementia | History of dementia based on registry data from the Finnish Care Register for Health Care (ICD-8: 290, ICD-9: 290–294, ICD-10: F00–06) |
| Prostate disorder | Use of medication for prostate disorders |
| Amyotrophic lateral sclerosis (ALS) | History of ALS based on register-data from the Finnish Care Register for Health Care (ICD-8: 348, ICD-9: 3352A, ICD-10: G122) |
| Parkinson’s disease | History of Parkinson’s disease based on registry data from the Finnish Care Register for Health Care (ICD-8: 342, ICD-9: 3320A, 3321A, 330A, ICD-10: G20–23) |

All data was retrieved prior to baseline. eGFR=estimated glomerular filtration rate

Supplemental Table S4. Prevalence of chronic conditions and risk of mortality

|  | **Prevalence, % (n)** | **Hazard ratio (HR) for mortality, age- and sex-adjusted** | **p-value** |
| --- | --- | --- | --- |
| ***Vascular comorbidities*** |  |  |  |
| Hypertension | 40.6 (1,653) | 4.6 (3.8–5.5) | <0.001 |
| Severe retinopathy | 35.1 (1,427) | 3.9 (3.3–4.6) | <0.001 |
| Diabetic kidney disease | 24.6 (1,002) | 6.4 (5.5–7.4) | <0.001 |
| Coronary heart disease | 5.9 (242) | 2.7 (2.2–3.3) | <0.001 |
| Peripheral vascular disease | 4.8 (195) | 5.1 (4.2–6.1) | <0.001 |
| Myocardial infarction | 3.3 (136) | 2.9 (2.3–3.6) | <0.001 |
| Stroke | 2.5 (101) | 3.6 (2.8–4.5) | <0.001 |
| Heart failure | 1.3 (52) | 5.3 (3.8–7.4) | <0.001 |
| ***Autoimmune diseases*** |  |  |  |
| Thyroid disease | 8.8 (358) | 0.9 (0.7–1.1) | 0.356 |
| Celiac disease | 2.2 (90) | 0.9 (0.5–1.6) | 0.833 |
| Rheumatoid arthritis | 2.1 (84) | 1.0 (0.7–1.6) | 0.885 |
| Addison’s disease | 0.2 (8) | 3.2 (1.0–10.1) | 0.043 |
| Multiple sclerosis | 0.2 (8) | 2.1 (0.7–6.6) | 0.193 |
| Atrophic gastritis | 0.1 (5) | 2.2 (0.5–8.7) | 0.271 |
| Myasthenia gravis | 0.0002 (1) | 56.4 (7.8–406.5) | <0.001 |
| ***Other diseases*** |  |  |  |
| Lung disease | 4.4 (179) | 1.1 (0.8–1.5) | 0.760 |
| Depression | 3.8 (156) | 2.1 (1.6–2.8) | <0.001 |
| Dyspepsia | 3.3 (134) | 3.4 (2.7–4.3) | <0.001 |
| Chronic pain | 3.0 (123) | 2.3 (1.7–3.1) | <0.001 |
| Alcohol abuse | 2.9 (120) | 3.0 (2.3–4.0) | <0.001 |
| Anxiety/insomnia | 2.6 (104) | 3.0 (2.2–3.9) | <0.001 |
| Epilepsy | 2.2 (91) | 1.2 (0.8–1.9) | 0.425 |
| Cancer | 1.2 (50) | 1.7 (1.1–2.6) | 0.015 |
| Glaucoma | 1.2 (48) | 1.5 (1.0–2.3) | 0.072 |
| Osteoporosis | 0.8 (34) | 3.8 (2.4–6.0) | <0.001 |
| Substance abuse | 0.5 (20) | 3.2 (1.6–6.1) | <0.001 |
| Schizophrenia | 0.5 (19) | 2.9 (1.4–6.1) | 0.005 |
| Dementia | 0.3 (12) | 3.0 (1.4–6.4) | 0.003 |
| Prostate disorder | 0.3 (11) | 1.9 (0.8–4.2) | 0.125 |
| Amyotrophic lateral sclerosis | 0.07 (3) | 1.6 (0.4–6.3) | 0.530 |
| Parkinson’s disease | 0.05 (2) | 44.3 (11.0–179.0) | <0.001 |

Data are hazard ratios with 95% confidence intervals.
